# A comparison of two forecasting models for COVID-19 hospitalizations using wastewater concentration data

**DOI:** 10.64898/2026.09.08.26362558

**Authors:** Alexander C. Keyel, Dustin. T. Hill, Kaitlyn E. Johnson, Daniel Lang, Eli S. Rosenberg, Kathleen Bush, David A. Larsen

**Affiliations:** Wadsworth Center, New York State Department of Health, Albany, NY, USA; Department of Public Health, Maxwell School of Citizenship and Public Affairs, Syracuse University, Syracuse, NY, 13244, USA; Department of Infectious Disease Epidemiology and Dynamics, London School of Hygiene and Tropical Medicine, London, UK; Center for Environmental Health, New York State Department of Health, Albany, NY, USA; Office of Science, New York State Department of Health, Albany, NY, USA

**Keywords:** COVID-19, epidemiological forecasting, GLMM, disease modelling, wastewater surveillance, wwinference

## Abstract

Wastewater surveillance has become a prominent part of public health efforts to track circulating pathogens and has been incorporated into disease forecasting models. We compared two recent wastewater forecasting models for forecasting COVID-19 hospitalizations using SARS-CoV-2 concentrations in wastewater: a generalized linear mixed model (GLMM) and a Bayesian mechanistic model known as wwinference. We retrospectively produced 1-week ahead forecasts using these two conceptually different models across 10 regions in New York state, from September 2022 – mid-April 2024. We compared the performance of forecasts produced from the two models fit with wastewater and clinical data to a version of each model fit to only clinical data. Models were scored against observations using Continuous Ranked Probability Scores for each forecast (n = 363 forecasts). Of the two models, the GLMM showed improved forecast performance across space and time when evaluated on a natural scale compared to the wwinference model, but no significant difference in forecast performance was found on a log-scale, which evaluates the relative rather than absolute error. Consequently, either model could be used to forecast COVID-19 hospitalizations. Including wastewater concentrations in these models at these spatial and temporal scales did not provide additional forecasting benefit beyond just clinical data. The performance of a wastewater-only model relative to simple null models; however, demonstrates that there is a clear forecasting signal present in wastewater, thus we think the lack of change is likely due to the strong signal from the clinical measures included in the model at this spatial and temporal scale.

## 1. Background

Wastewater surveillance for infectious disease has a long history [1] and has undergone a recent expansion through the National Wastewater Surveillance System in the United States, in addition to expanding globally. Wastewater surveillance offers several advantages over other surveillance methods, as it provides a non-invasive means to survey large portions of the population, and may serve as an early warning signal for disease presence or expansion [2]. It works on the principle that an infected individual sheds pathogens, pathogen genetic material, and/or other pathogen derivatives into the wastewater system [3], which can then be detected or quantified in a laboratory from wastewater samples. An early warning signal from wastewater is expected in part due to the shedding of pathogens by infected people who are not yet symptomatic or have not yet become infected severely enough to be admitted to the hospital [4]. In contrast, alternative approaches to forecasting disease burden often rely upon clinical cases, test positivity, or hospitalization data [5], thus potentially leading to delayed predictions relative to those provided by wastewater. Changes in testing and reporting rates may also change reported clinical case numbers but are not expected to change wastewater measurements. Similarly, hospitalization will only capture the most severe cases. However, wastewater is expected to reflect all individuals contributing to a catchment area, while asymptomatic individuals and unreported cases (e.g., due to at-home testing) will not be included in incidence estimates from clinical cases.

SARS-CoV-2 provides an excellent case study for evaluating the capacity of wastewater surveillance to contribute to infectious disease forecasts. SARS-CoV-2 has resulted in more than one million deaths in the United States and more than seven million reported deaths worldwide [6]. It continues to hospitalize individuals across the globe. Wastewater surveillance was quickly adapted to SARS-CoV-2 shortly after its emergence [7]. While a variety of surveillance methods for SARS-CoV-2 exist [5], numerous studies have demonstrated a strong correlation between the amount of SARS-CoV-2 RNA in wastewater and human cases and/or hospitalizations [8–11]. The amount of SARS-CoV-2 RNA is expected to correlate with incidence [12], and hospitalizations [13]. Hospital admissions are generally considered to represent a reliable indicator of COVID-19 incidence and are routinely reported in public health surveillance, though not all jurisdictions report their data publicly. Furthermore, patterns in COVID-19 hospitalizations may be of particular interest for hospital management [14].

The New York Wastewater surveillance program provides an excellent dataset to compare model forecasts, as prior research has shown a strong relationship between hospitalizations and SARS-CoV-2 wastewater concentrations [15], has had a wastewater surveillance network in place since 2020, and includes urban, suburban, and rural sewersheds.

We compared one-week ahead forecasts generated by two recent hospitalization forecasting models [15, 16] that can incorporate SARS-CoV-2 concentrations in the prediction process. We compared the models to each other, parameterizing them with and without wastewater, to determine the strengths and weaknesses of each model, and to identify the degree to which wastewater improved model forecasts. We also examined whether there was regional variation in model performance. Each modeling method was fit from clinical data only (models without wastewater) and then with clinical data and wastewater data added (models with wastewater), enabling a comparison of four different forecast outputs: i.e. 2 models × 2 data sources (clinical data only and clinical plus wastewater, Fig. S1). Additionally, a wastewater-only model was fit, and two null models were considered to provide a baseline for model skill. Model forecasts were evaluated with the Continuous Ranked Probability Score (CRPS), and scores were statistically compared across forecast dates and regions to identify statistically significant differences in forecast performance, considering base model, whether or not wastewater was included, and the interaction between base model and wastewater.

## 2. Methods

### 2.1 Data set description

The first reported COVID-19 case in NYS occurred on 29 February 2020. Wastewater surveillance in New York State began in April 2020, but samples from a routine statewide network (every county) began in August 2022 [17]. For this study, we restricted the data set to September 2022 through mid-April 2024 [18], to ensure a robust wastewater data set for the comparison. The analysis presented here is based on 363 forecasts distributed evenly throughout the study period, with the same dates selected for each of the 10 regions (Figs. S2, S3). All dates two weeks apart that had sufficient wastewater data for both models were included as forecast dates. Surveillance ranged from a minimum of 8 wastewater treatment plants contributing to a forecast, to a maximum of 38 wastewater treatment plants within a region (Fig. S4a). Most forecasts included >100 samples total (Fig. S4b) and >10 samples per wastewater treatment plant in the forecast (Fig. S4c.). We required at least one wastewater sample 80 or more days prior to the forecast. Hospitalization data were from the NYS Health Electronic Response Data System (HERDS) [19].

Hospitalizations were aggregated across all hospitals within a region. Hospitalizations were reported with daily resolution (except for two dates) to the state until 15 April 2022. Starting April 15, hospitalizations were no longer reported on weekends or holidays, with all accumulated hospitalizations reported on the next business day. We backfilled these accumulated hospitalizations to fill the gaps in the data set. (e.g., if 30 hospitalizations were reported on a Monday, and there was no report for the previous Saturday/Sunday, each day would be assigned 10 hospitalizations). We aggregated hospitalizations to a weekly aggregate as our forecast input to accommodate changes in NYS hospitalization data reporting. The hospitalization data set reported out all individuals hospitalized with COVID-19. As the wwinference model requires daily data, we then distributed each week’s hospitalizations over the individual days within the week. Reported SARS-CoV-2 concentrations (gene copies per mL) were used as the model input, as prior exploratory analysis showed no strong benefits of normalizing by Pepper Mild Mottle virus, crAssphage, or flow rate. This is consistent with studies in the literature, some of which have shown no benefit or inconsistent benefits of normalization [20, 21]. Methods to quantify the amount of SARS-CoV-2 RNA from wastewater samples varied by region with most regions processed by a lab using ultracentrifugation followed by RT-qPCR [22] and New York City using PEG precipitation and RT-qPCR initially before switching to dPCR [23, 24]. The switch to dPCR on March 12, 2023 led to an increase in measured concentrations, and a linear transformation was used based on the multiplicative factor of each sampling site’s ratio of RT-qPCR results to dPCR results from concurrent quantifications for the two methods to adjust the data set to be on the same scale. For example, if one site had dPCR results that were 15 times higher, the RT-qPCR data were multiplied by 15. There was also a second methods change for New York City on March 3, 2024, but changes in observed PCR values were not as large, so no adjustment was made.

### 2.2 Forecasting Models

#### 2.2.1 wwinference model

The wwinference model [16, 25] is a hierarchical semi-mechanistic renewal model that jointly infers infection dynamics from a “global” count dataset (in this case, hospital admissions) and wastewater concentrations from individual wastewater treatment plants. The model works by estimating the effective reproductive number (Rt) in each subpopulation (where subpopulations are defined by the wastewater catchment areas and the remaining subpopulation), modeled as deviations from a central infection dynamic. The renewal equation is used to estimate latent incident infections in each subpopulation, which are then used to generate the expected observations via convolutions with the corresponding delay distributions. For the hospital admissions, the total incident infections are estimated as a sum of the subpopulation incident infections and then convolved with the delay distribution from incident infection to hospital admission and scaled by the infection to hospital admissions rate. The observed hospital admissions are assumed to follow a negative binomial observation model with an inferred day of the week effect. For wastewater concentrations, a triangular function in log scale describes the shedding kinetics delay distribution, and this is scaled by the average number of genomes shed per infection and divided by the volume of wastewater produced per person per day. The observed wastewater concentrations are assumed to follow a lognormal observation model. The model is fully Bayesian and fit using Hamiltonian Monte Carlo implemented in R [27] and the probabilistic programming language Stan [28] using the cmdstanr package [29]. For more information on the model and implementation, see the wwinference package documentation and model definition [16, 25]. We used version 0.1.1. We used the most recent 90 days of hospitalizations to fit the wwinference model.

#### 2.2.2 GLMM forecasting model

The second model was a generalized linear mixed model (GLMM) that was fit to estimate new hospital admissions per 100,000 population (hospital incidence) for each region and has been described previously [22]. The model uses a Gaussian distribution and an autoregressive error structure on the date (each week) to correct for repeated measures from each region. The model structure is defined in Equation 1.

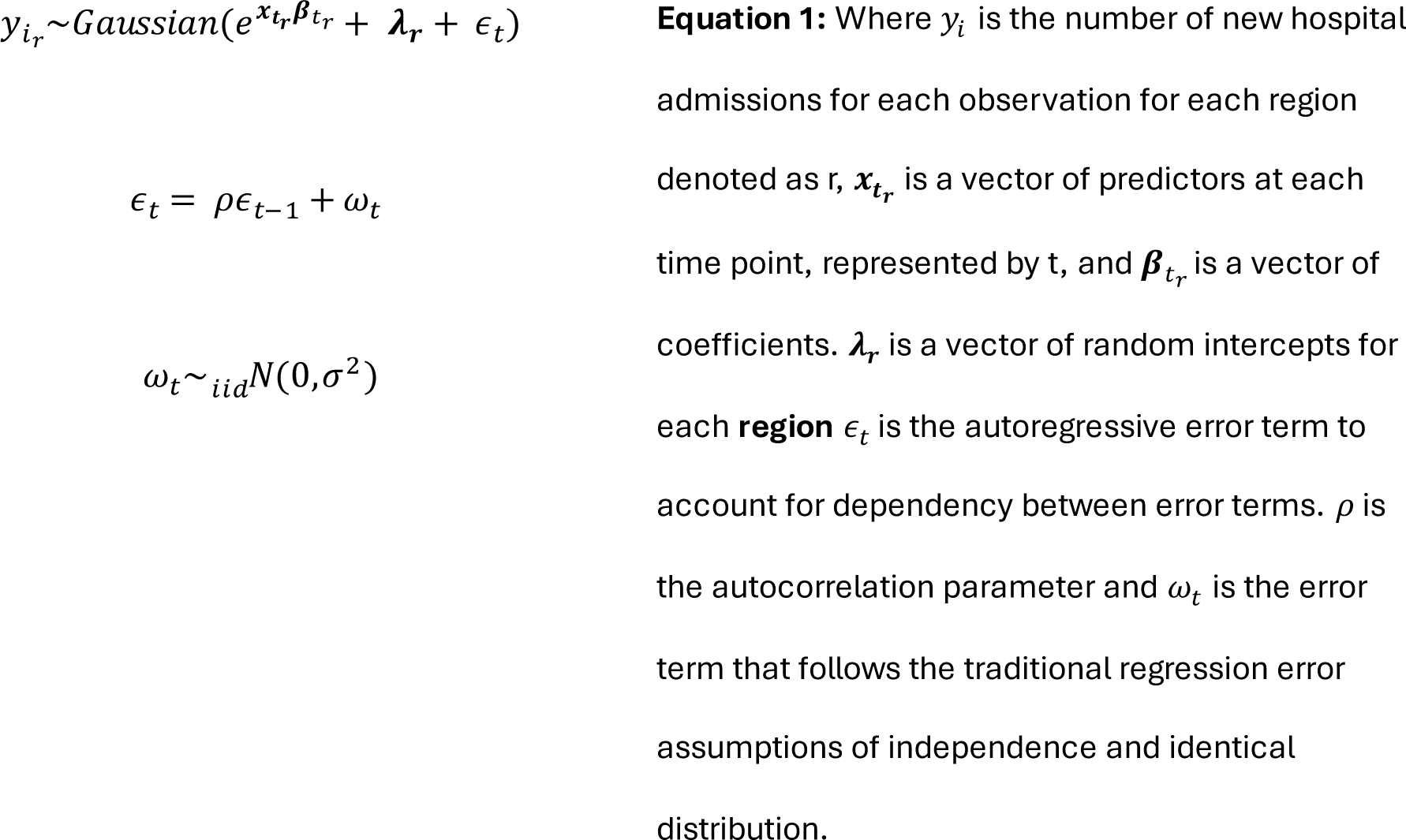

The model predicts hospital incidence using three covariates: log-transformed weekly SARS-CoV-2 average concentration in gene copies per mL from the previous week, COVID-19 hospital incidence from the previous week, and the square-root transformation of new COVID-19 cases from the previous week. Square-root transformation was applied to the case data to avoid collinearity with SARS-CoV-2 wastewater concentration. Hospital incidence per week is derived from the sum of hospital admissions in each week as reported by NYS [19] and then transformed into incidence per 100,000 population by region. Case data per week are derived from the sum of new cases reported to NYS [30]. SARS-CoV-2 concentrations are collected on different sampling days each week at each site. To get weekly values, the wastewater data were transformed into a daily time series using linear interpolation between sample days. Gaps larger than eight days were not filled since nearly all sites sample once weekly. Then a seven day right-adjusted rolling average was calculated on the daily data. Last, the data were aggregated to the regional level per week using a population weight of the sampling site’s population. The R package and function “glmmTMB” was used for model fitting [31]. Data were restricted to the most recent 24 weeks to allow the model to account for any changes in the relationship between hospitalizations and wastewater concentration over time. The GLMM model was also fit without wastewater as a predictor, to assess the predictive skill of the clinical metrics alone.

#### 2.2.3 Wastewater-only Model

Additionally, the GLMM model was fit as above, but omitting the clinical data, to demonstrate the predictive skill of wastewater in the absence of clinical data.

#### 2.2.4 Baseline Null models

For baseline models, we chose a model that predicted the median value to be the current hospitalization count. A prediction interval was constructed by evaluating the error from the historical variation in the observations relative to the prior week and calculating the standard deviation of those errors. A second baseline model took the previous 3 observations and fit a line to them using simple linear regression. This provided us with two imperfect baseline models – one expected to perform poorly for sustained increases/decreases, the other expected to perform poorly for a change in direction of hospitalizations during the forecast period.

#### 2.3.1 Forecasting Model Comparison

Retrospective 1-week ahead forecasts (n = 363) for the following week’s COVID-19 hospitalizations were generated for each of the forecasting models. The forecast dates were generated, spaced two weeks apart, for each of 10 economic development regions in NYS (Fig. S2) for the entire timeframe of the study (Fig. S3). Two weeks was selected to provide some measure of independence. A 1-week ahead forecast was chosen because 1) this represents the next time step for a weekly forecast, 2) it most closely matched the 10-day optimal performance of the GLMM model based on daily data, and 3) reduces the potential for model results to diverge from observations over time (representing a best-case forecast for the models). Forecasts were compared to observed hospitalizations per 100,000 population. Reporting followed the EpiForge guidelines [32] (Table S1).

#### 2.3.2 Forecast Scoring

Model forecasts were scored using CRPS [33, 34]. CRPS is a proper scoring method that can be used to incorporate forecast uncertainty directly into the scoring process. CRPS is a generalization of the mean absolute error (MAE) for probabilistic forecasts. Similar to the MAE, larger CRPS scores represent less skillful predictions, and similar to the MAE, the CRPS scales with the magnitude of the observations/forecasts (e.g. forecasts for higher incidence will typically have higher CRPS just as the absolute error will typically be higher when the magnitude is higher). CRPS are continuous, not binary, giving closer predictions a better score than further predictions (unlike other scoring methods which may use a binary classification into accurate and inaccurate).

CRPS were evaluated in two ways: 1) using the untransformed observations (natural scale) and by using log-transformed observations and predictions (log scale). While a natural scale may provide a more intuitive result, evaluations on the log scale put errors into the context of the growth rate rather than the absolute number [35]. For example, being off by 1 represents a 50% error if there are 2 cases, but a 1% error if there are 100 cases.

CRPS was computed from model predictions and observations using the scoringRules package [34]. The scoringRules package can take either a distribution of predictions (such as generated by wwinference), or a mean and standard deviation (as generated by the GLMM model). For the log-scale evaluation of the GLMM model, samples were drawn from the distribution described by the mean and standard deviation on a natural scale, and then those were log-transformed and entered into scoringRules, as log-transforming the mean and standard deviation would not have preserved the statistical distribution of the predictions.

#### 2.3.3 Statistical Comparison of Forecast Scores

The CRPS were then statistically compared. First, each forecast date for each region was treated as an independent data point. Models were compared across dates and regions using paired t-tests to compare model pairs. This has the strength of providing a simple, interpretable statistical test that controls for differences in SARS-CoV-2 variant, time, pandemic trend, or magnitude of hospitalizations, as the difference in CRPS between forecasts was the statistic of interest. Nocorrection was madefor multiple comparisons, because each pairwise comparison (between model structure and/or input data) presented was of interest on its own, and correcting for multiple comparisons would have reduced the statistical power of these comparisons.

Second, as wwinference and the GLMM models were of primary interest, a second analysis was conducted using a multipleregression framework limited to those models’ CRPS scores (Equation 2). This analysis tested for differences in model, presence or absence of wastewater, and region, with a model × region interaction term to test whether the difference between models varied by region. Specifically, the goal was to determine if the top model varied by region (e.g., one model performing better in rural areas, and the other performing better in populated areas). This analysis used the CRPS from individual forecast dates (i) for each region (j) as independent inputs, and did not account for the matched structure within each forecast-region prediction.

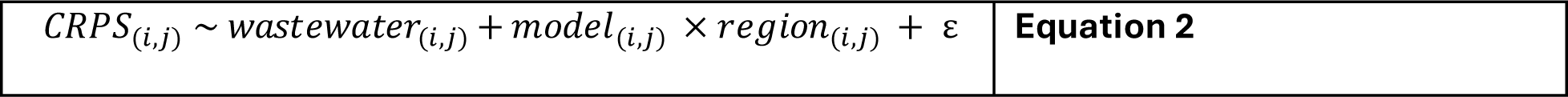

Analyses were conducted separately for CRPS scores evaluated on a natural scale and evaluated on a log scale. All statistical analyses were conducted in R [27].

## 3. Results

The GLMM model represents a statistical fitting approach, while the wwinference model is a Bayesian semi-mechanistic renewal-based model. The GLMM model runs relatively quickly (seconds), while the wwinference model was more computationally intensive (minutes to hours), depending on how many wastewater treatment plants and samples were included.

When evaluated across regions and across the time period, no significant difference was observed between either model with wastewater and the same model without wastewater (Table 1, paired t-tests, all p ≥ 0.45 with 362 df). On a natural scale, the GLMM model outperformed the wwinference model when both were fit without wastewater (Table 1, difference -0.063, p = 0.02) and when they were fit with wastewater (Table 1, difference -0.062, p = 0.02). On a log scale, there was no significant difference between the two models without wastewater (Table 1, difference = 0.002, p = 0.58) or with wastewater (Table 1, difference = 0.003, p = 0.44). Both the wwinference model and GLMM model showed increased forecast skill compared to two simple null models on both the natural scale and the log scale (Table 1, all p <0.001).

**Table 1.**
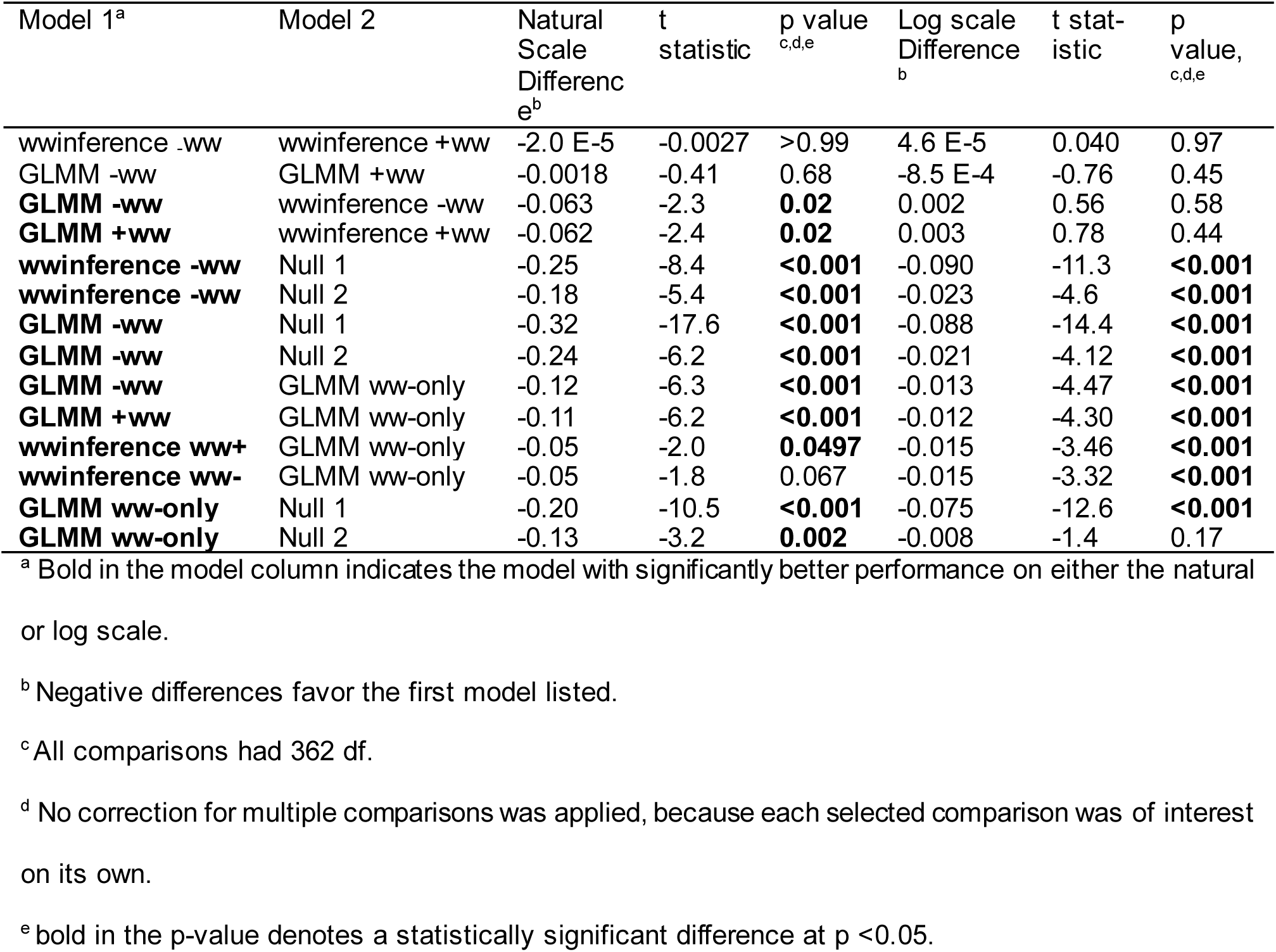
Paired t-test results for selected comparisons. “-ww” refers to the model without wastewater, and “+ww” refers to the model with wastewater.

| Model 1<sup>a</sup> | Model 2 | Natural Scale Difference<sup>e</sup> | t statistic | p value<sup>c,d,e</sup> | Log scale Difference<sup>b</sup> | t statistic | p value, <sup>c,d,e</sup> |
| --- | --- | --- | --- | --- | --- | --- | --- |
| wwinference -ww | wwinference +ww | -2.0 E-5 | -0.0027 | >0.99 | 4.6 E-5 | 0.040 | 0.97 |
| GLMM -ww | GLMM +ww | -0.0018 | -0.41 | 0.68 | -8.5 E-4 | -0.76 | 0.45 |
| **GLMM -ww** | wwinference -ww | -0.063 | -2.3 | **0.02** | 0.002 | 0.56 | 0.58 |
| **GLMM +ww** | wwinference +ww | -0.062 | -2.4 | **0.02** | 0.003 | 0.78 | 0.44 |
| **wwinference -ww** | Null 1 | -0.25 | -8.4 | **<0.001** | -0.090 | -11.3 | **<0.001** |
| **wwinference -ww** | Null 2 | -0.18 | -5.4 | **<0.001** | -0.023 | -4.6 | **<0.001** |
| **GLMM -ww** | Null 1 | -0.32 | -17.6 | **<0.001** | -0.088 | -14.4 | **<0.001** |
| **GLMM -ww** | Null 2 | -0.24 | -6.2 | **<0.001** | -0.021 | -4.12 | **<0.001** |
| **GLMM -ww** | GLMM ww-only | -0.12 | -6.3 | **<0.001** | -0.013 | -4.47 | **<0.001** |
| **GLMM +ww** | GLMM ww-only | -0.11 | -6.2 | **<0.001** | -0.012 | -4.30 | **<0.001** |
| **wwinference ww+** | GLMM ww-only | -0.05 | -2.0 | **0.0497** | -0.015 | -3.46 | **<0.001** |
| **wwinference ww-** | GLMM ww-only | -0.05 | -1.8 | 0.067 | -0.015 | -3.32 | **<0.001** |
| **GLMM ww-only** | Null 1 | -0.20 | -10.5 | **<0.001** | -0.075 | -12.6 | **<0.001** |
| **GLMM ww-only** | Null 2 | -0.13 | -3.2 | **0.002** | -0.008 | -1.4 | 0.17 |

<sup>a</sup> Bold in the model column indicates the model with significantly better performance on either the natural or log scale.
<sup>b</sup> Negative differences favor the first model listed.
<sup>c</sup> All comparisons had 362 df.
<sup>d</sup> No correction for multiple comparisons was applied, because each selected comparison was of interest on its own.
<sup>e</sup> bold in the p-value denotes a statistically significant difference at $p < 0.05$ .

A wastewater-only GLMM mode had significantly higher CRPS (decreased forecast performance) than the GLMM models that included clinical data on the natural scale (Table 1, difference = -0.12, p <0.001) and on the log scale (Table 1, difference =-0.013, p <0.001). The wastewater-only GLMM also had significantly higher CRPS on the log-scale compared to the wwinference model (all p <0.001), but on a natural scale, the difference was borderline significant, or borderline non-significant, depending on the comparison (wwinference with wastewater, difference = -0.05, p = 0.0497, wwinference without wastewater, difference = -0.05, p = 0.067, Table 1).

**Fig. 1.**
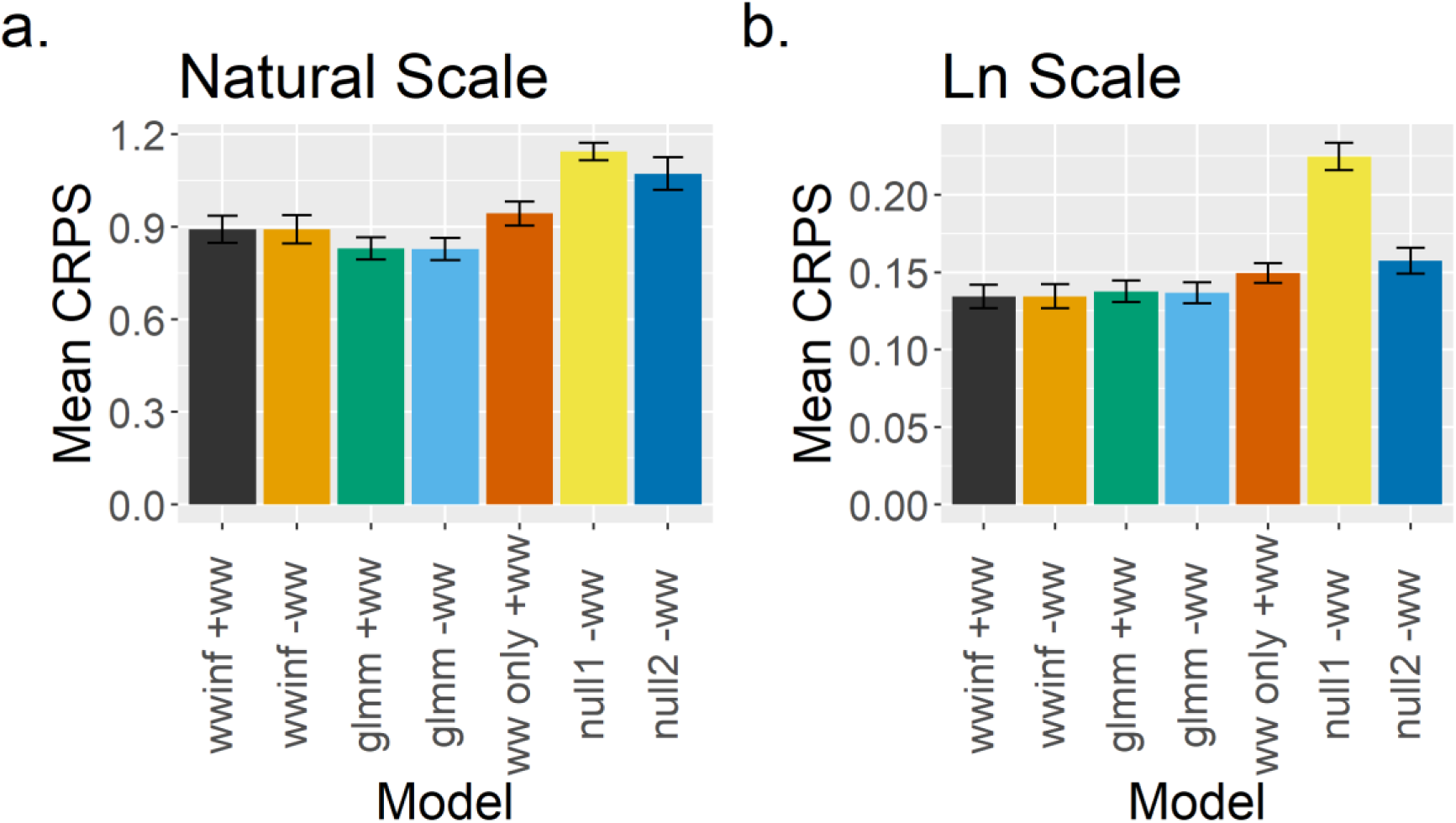
Mean CRPS scores across all forecasts and regions for seven models. Models are the wastewater inference model (wwinf) and the generalized linear mixed-models model (glmm) with (+) and without (-) wastewater when compared across locations and forecast dates, a glmm wastewater only model (ww only +ww) and a persistence null model (null1) and a simple linear trend null model (null2). Evaluations were performed on (a) a natural and (b) log scale. Colors correspond to the different models. Error bars indicate ±1 standard error. Note that statistical tests were conducted on paired differences and not a direct comparison of overlap in standard errors.

CRPS scores varied by region (Fig. 2). Model forecasts were more skillful for New York City, the region with the lowest (best) CRPS score on a natural scale, than for Central New York, Mohawk Valley, North Country, and the Southern Tier (Table S2) when evaluated on a natural scale. On a log scale, the Finger Lakes region had the lowest CRPS score, and showed significantly improved skill relative to Central New York, Mohawk Valley, North Country, and the Southern Tier (Table S3). There were not strong regional differences between models when evaluated on a natural scale (regional differences in model performance: -0.10 to 0.26, all p > 0.13; see interaction terms in Table S2). On a log-scale, there was a near-significant improvement for the wwinference model relative to the glmm model for New York City (Table S3, p = 0.09, all other region interaction terms p > 0.5).

**Fig. 2.**
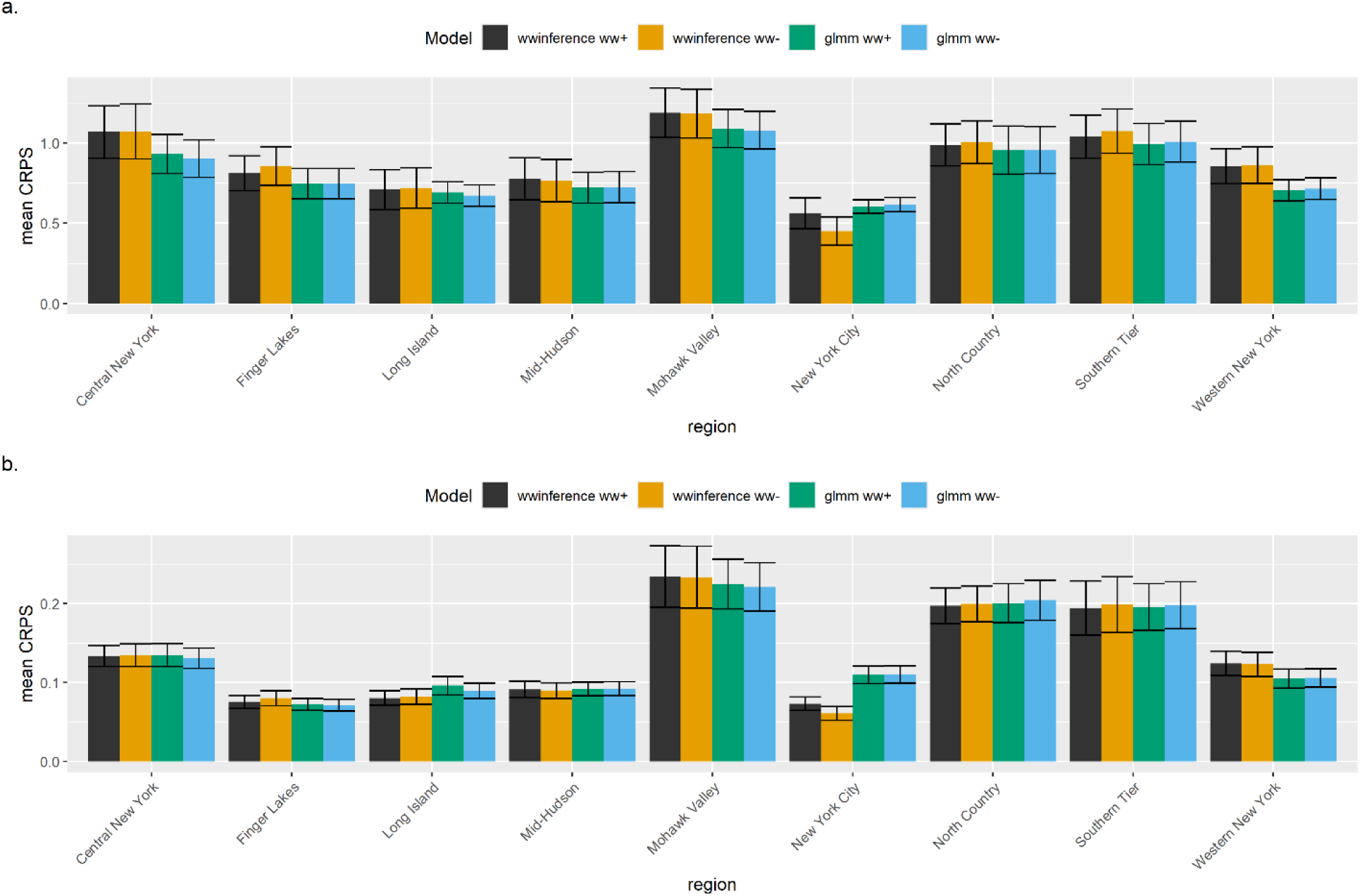
CRPS scores by region for hospitalizations per 100,000 population on a) natural scale and b) on a log scale. ww+ indicates models fit with wastewater, and ww-indicates models fit without wastewater.

While the GLMM model performed better on a natural scale (no difference on a log-scale), there was no clear pattern in the timing of when it was performing better (Fig. 3a, b). For the Long Island example, the largest errors appear to correspond to peaks in hospitalizations (Fig. 3d, Fig. 4). In the selected example (Fig. 4), predictions were higher than hospitalizations on October 8, 2023 (wwinference CRPS = 2.3, GLMM CRPS = 1.0) and on January 14, 2023 (wwinference CRPS = 4.8, GLMM CRPS = 2.2). There was no pattern for either model for when including wastewater improved (or reduced) forecast performance (Fig. 3c). Examining the models closely showed many values close to a 1:1 line for a plot of predictions without wastewater on the x axis and predictions with wastewater on the y axis (Fig. 5), suggesting close predictive ability of both pairs of models.

**Fig. 3.**
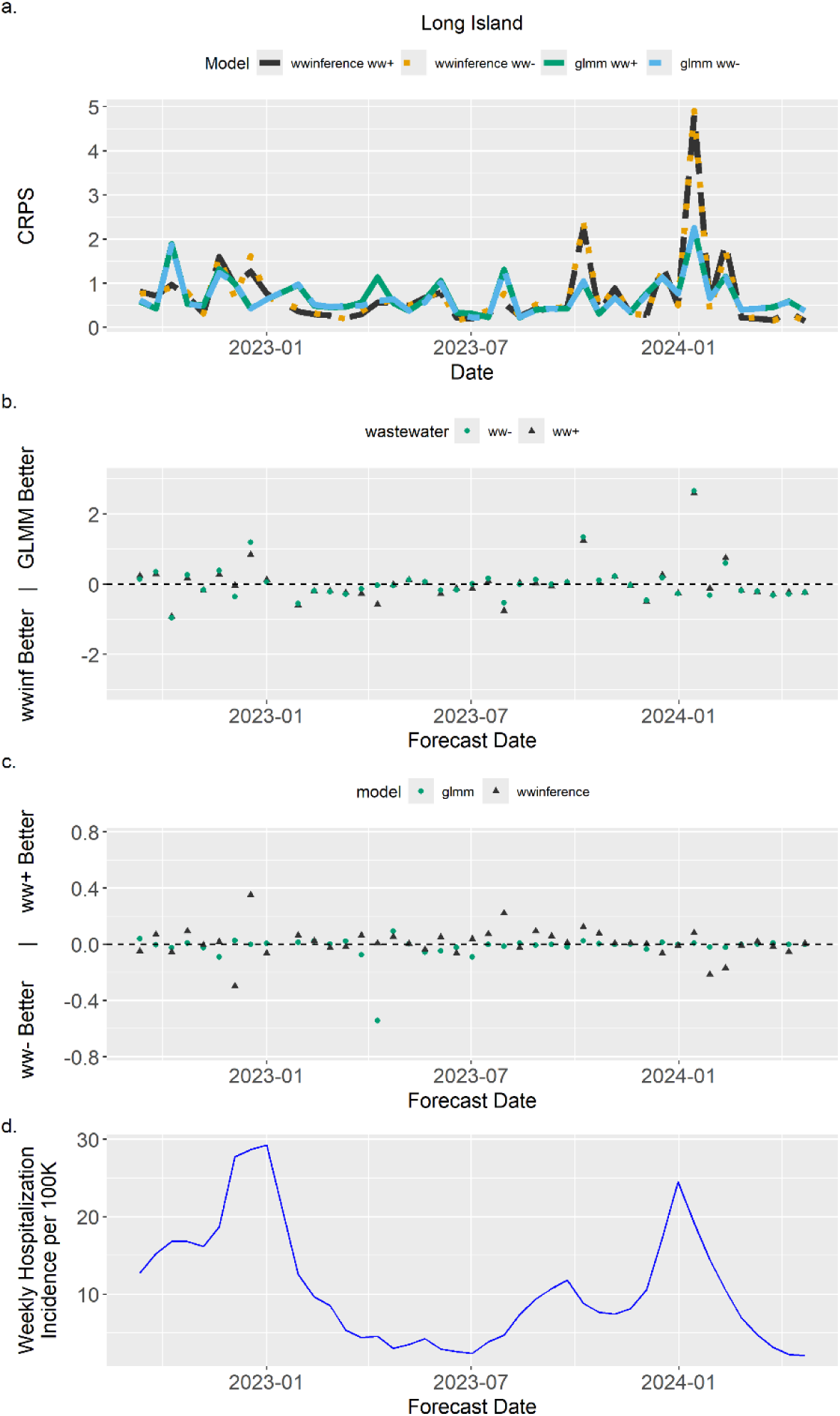
CRPS scores evaluated on natural scale, using the Long Island region as an example to show a) CRPS scores over time and b) difference in CRPS between models (wwinference model – glmm model) for models with and without wastewater, c) difference in CRPS by model type comparing performance of models with and without wastewater (model without wastewater – model with wastewater), and d) weekly hospitalization incidence per 100,000 population for context. ww+ indicates models fit with wastewater, and ww-indicates models fit without wastewater.

**Fig. 4.**
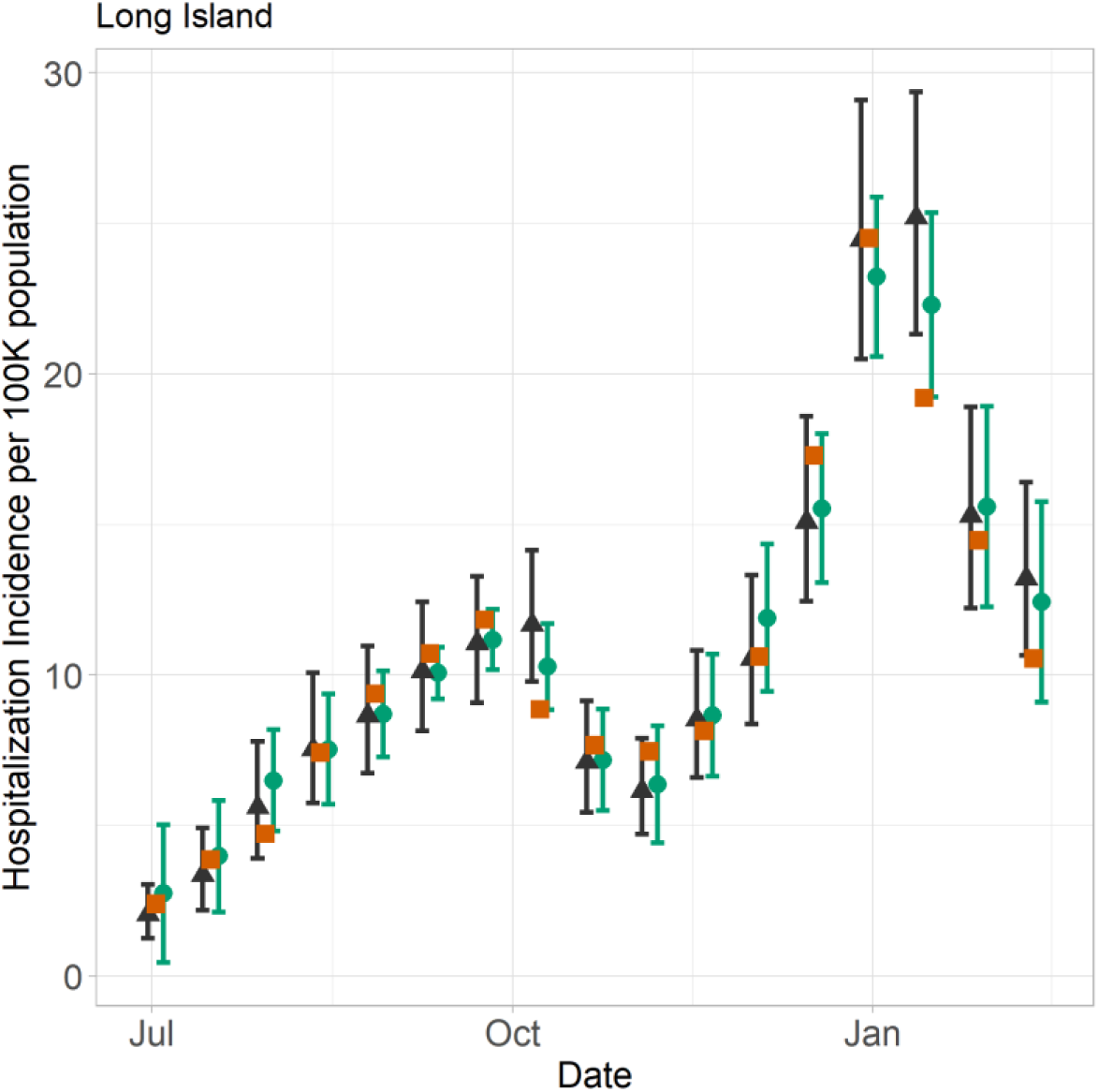
Natural scale observations (orange square) for July 1, 2023 – Feb 15 2024, for the Long Island Region compared to model predictions: wwinference with wastewater (dark triangle) and GLMM with wastewater (green circle). Model predictions without wastewater not shown. Error bars show 95% prediction interval. Hindcast predictions were jittered for visibility. Date range was selected to illustrate a rise and fall in COVID-19 hospitalizations for a single region.

**Fig. 5.**
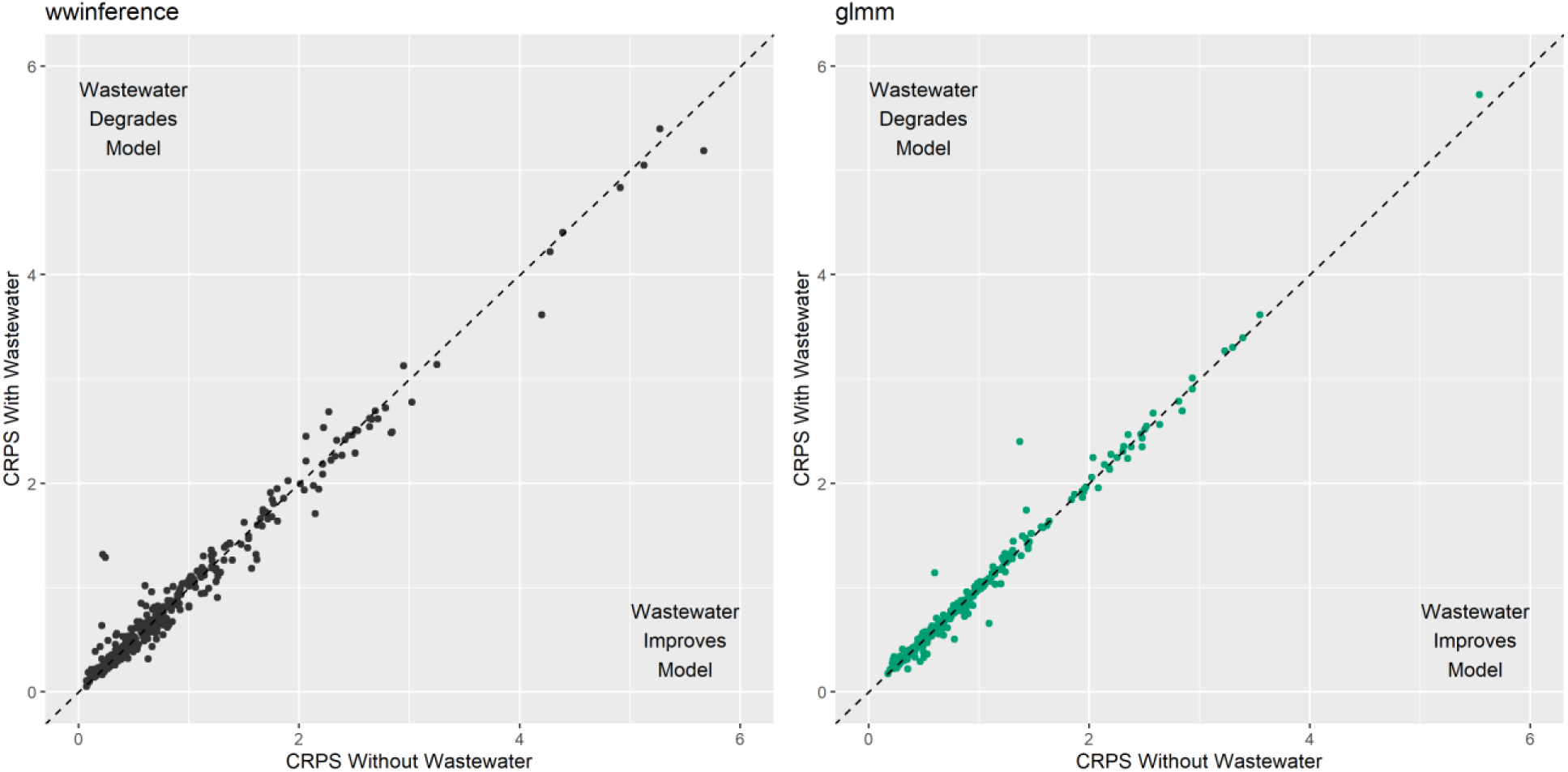
Continuous Ranked Probability Score (CRPS) for a) the wwinference and b) glmm models with (y -axis) and without (x-axis) wastewater when CRPS is evaluated on a natural scale. Deviations below the dashed 1:1 indicate model forecasts with increased skill with wastewater, while points above the 1:1 line indicate decreased model skill with inclusion of wastewater.

## 4. Discussion

We evaluated the performance of two different forecasting models with and without wastewater using a probabilistic scoring metric that incorporates the uncertainty in the forecast distribution, across one state wastewater surveillance system over multiple years. We found that the wwinference model and the GLMM performed equally well, both scoring significantly better than the null models regarding CRPS in both the natural and log-scales, with and without wastewater. Consequently, either model could be used for hospitalization forecasting, but the GLMM model may be easier to set-up and faster to run for public health forecasting, as it does not have a Stan dependency, and the Monte Carlo simulation can be time-consuming. The GLMM model also showed improved performance on a natural scale, but not on a log scale (Fig. 1, Table 1). This suggests that there may have been some improvement in forecasting at higher incidence, but any such improvement was small relative to the total incidence. Both models produced forecasts that were accurate (within ± 1 CRPS on a natural scale and ± 0.2 CRPS on a log scale, Fig. 3b,c) the majority of the time, however, each model also produced forecasts that deviated from the observed COVID-19 hospitalizations, as can be seen in Figs. 3 and 4. We are not the first to observe this with prior studies showing that even models built on case data struggled to produce accurate forecasts for COVID-19 hospitalizations [36]. The model forecasts were not identical, despite no significant overall difference in performance between the models across the study period. Future investigation could examine the points at which each model deviated, especially in the cases of the larger model errors, where these deviations may provide insights into ways to improve model performance.

In this study, a strong, additive signal from wastewater was not observed for either model. This result is not unique in the literature [25, 37], but does differ from prior publications that have shown an additive signal [15]. We note that there were different modeling choices between this study and Hill et al. 2023, including the use of a different hospitalizations dataset, the choice of time period analyzed, and the spatial scale of the modeling unit (e.g., region vs. county spatial resolution, weekly vs. daily predictions). The Modifiable Areal Unit Problem is a well-known pattern from ecology, where patterns at one spatial or temporal scale may not be the same as at another spatial or temporal scale [38–40]. Consequently, we hypothesize that these differences are scale-dependent, and that in the present study, the aggregated hospitalization data provided a stronger signal that overlapped with the skill provided by the strong wastewater signal. This is a topic warranting further research. This also highlights the importance of accurate and timely hospitalization reporting for disease forecasting. In NYS, COVID-19 hospitalization data are currently available on a weekly basis, while wastewater surveillance data are available within 3-5 days of a sample being collected.

Another explanation for the lack of an additive signal is that wastewater concentrations may be a reliable indicator of infection trends but could differ from hospitalizations if transmission patterns for vulnerable populations differ from those of the general public. COVID-19 hospitalization risk mitigation behaviors significantly differ by age group [41] and vaccination rates also differ among adults 18+ and those 65+ [e.g., 42]. Consequently, community transmission as measured in wastewater may not fully capture transmission among the most vulnerable individuals likely to be hospitalized. Further, hospitalizations are a rare event that occurs under specific circumstances and may not be as good an indicator of community transmission as test positivity or positive test incidence. Also, while we found that the wastewater only models were different in their predictive ability with reduced accuracy, we did not assess if they were meaningfully different from the perspective of interpretation of the forecasts. On both the natural and the log scale, the difference in CRPS was small. The difference was statistically significant, but future studies focused on implementation of forecasting might look at whether these differences matter practically. Last, our study assumes that wastewater and hospitalization data will always be reported at the same time. For this study, we assumed hospitalizations and wastewater concentrations were available through the day prior to the forecast. This will vary by location, and our findings suggest that either data source on their own provides greater forecast performance relative to simple clinical-based null models.

Potential limitations include choice of data set. This study used daily data aggregated to a weekly format, due to a change in data reporting to reporting weekly data in NYS HERDS data. Data processing after a certain point, and these were interpolated across all missing days). Daily resolution is available with the Statewide Planning and Research Cooperative System (SPARCS) data set; however, these data are currently only available after a 6-month lag. The choice of model could have also affected the outcome, although Rademacher et al. [37] considered five different models at regional and national scales and did not see forecast improvement by including wastewater in the models. It is also important to note that this study aimed at a technical comparison of the two forecasting models and their accuracy and did not assess whether the observed model accuracy was sufficient for decision-making with respect to COVID-19. Local health departments in New York State reported that hospitalization forecasts from wastewater have been useful throughout the COVID-19 pandemic [43]. This research was conducted across a range of urban, suburban, and rural locations in a temperate climate, so the results may be applicable to similar locations. However, changes in the input data’s spatial and temporal resolution and forecast window may lead to different outcomes. For example, evaluation on a log-scale and a natural scale led to different relative rankings of the GLMM and the wwinference model, and the near-significant difference for the New York City region suggests that region-specific variation may exist.

## 5 Conclusions

Wastewater surveillance has the potential to improve public health decision-making, situational awareness, and disease forecasting. Our systematic evaluation of two wastewater-based models designed to forecast hospitalizations revealed that the models performed similarly to each other, and to versions of the models without wastewater. If computation time and software setup are factors, the GLMM model is easier to implement and just as accurate as wwinference.

## List of abbreviations

CRPS: Continuous Ranked Probability Score
dPCR: Digital Polymerase Chain Reaction
GLMM: Generalized Linear Mixed Model
HERDS: Health Electronic Response Data System
MAE: Mean Absolute Error
NYS: New York State

## Declarations

### Ethics approval and consent to participate

Not applicable

### Consent for publication

Not applicable.

### Availability of data and materials

Hospitalization data are available online at https://coronavirus.health.ny.gov/daily-hospitalization-summary [19]. The majority of New York State wastewater data are publicly available [18], although the data set used here contains data from a few additional treatment plants that are not publicly available due to small population sizes of the sewershed catchments. Code available upon reasonable request. Tables of model CRPS scores by forecast date are included as supplemental materials, see Table S4 for a data dictionary.

## Authors’ contributions

Conceptualization: ACK, DTH; Methodology: ACK, DTH, KJ; Software: ACK, DTH, KJ; Validation: ACK; Formal Analysis: ACK; Investigation: ACK, DTH; Resources: KB, DL, DAL; Data Curation: ACK, DTH; Writing – original draft: ACK, DTH, KJ, Writing – Review & Editing: ACK, DTH, KJ, DL, ER, KB, DAL, Visualization: ACK, KJ; Supervision: DL, ER, KB, DAL, Project Administration ACK; Funding Acquisition: KB, DL, DAL.

## Acknowledgements

The authors would like to thank the New York State Wastewater Surveillance Network including Kathleen McDonough, Kirsten St. George, Chris Dunham, P. Bryon Backenson, Bridget Anderson, Mohammed Alazawi, Haley Kappus-Kron, Spencer Bruce, Isaac Ghinai, Kayla English, Dylan Morris and the McDonough Lab. The authors thank the PCR labs from Quadrant Biosciences, Ian M. Bradley, and Yinyin Ye from the University at Buffalo, Department of Civil, Structural and Environmental Engineering, Christopher Gobler, and Mian Wang from Stony Brook University, Karen Schmidt and Brenden Bedard at Genesee and Orleans County Health Department, and the New York City Department of Health.

## Supporting Information for

**Fig. S1.**
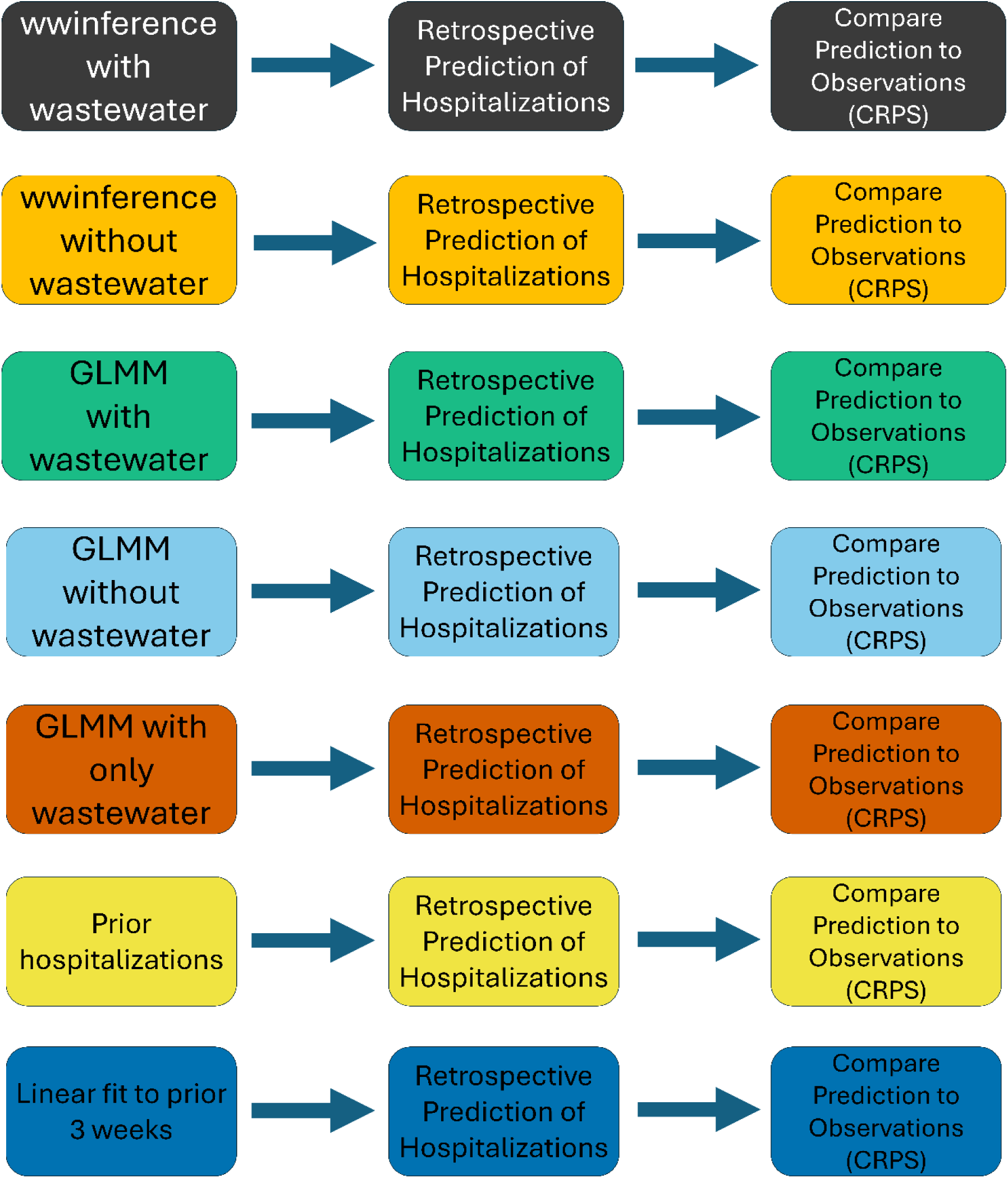
A schematic showing the four model outputs to be compared, along with two null models and a wastewater-only model to provide context for the models’ forecasting skill. Each of two models were run with and without wastewater, leading to four combinations. These retrospective forecasts were evaluated in a probabilistic framework, using the continuous ranked probability score from the scoringRules package in R. The GLMM model was also fit to only wastewater (no hospitalizations) to demonstrate the predictive capacity of wastewater in the absence of hospitalization data, and two simple models (current hospitalizations carried forward, and a linear fit to the three most recent weekly hospitalizations) were included to demonstrate that fitted models had predictive skill beyond simple null models.

**Fig. S2.**
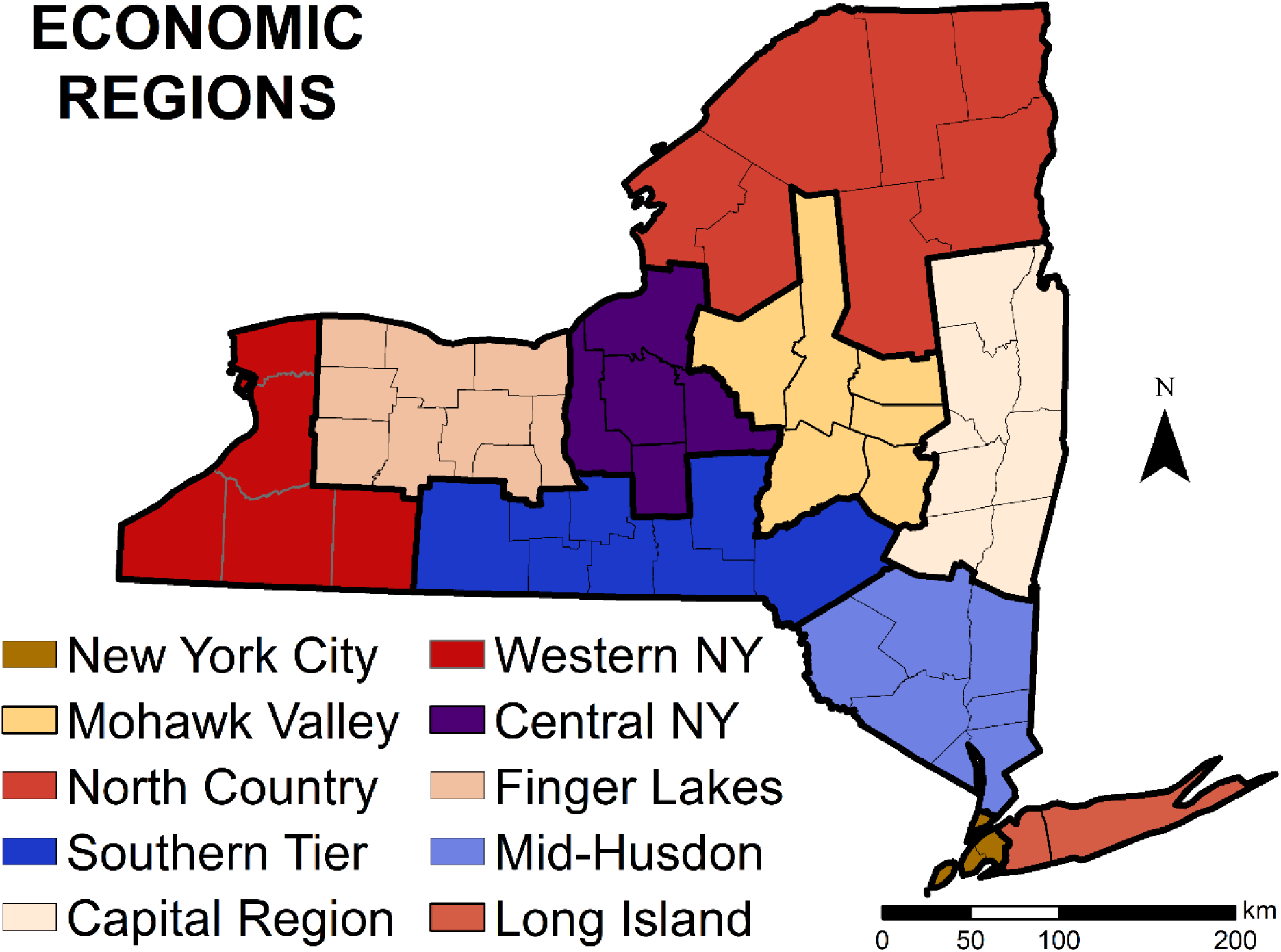
New York State and the 10 economic regions used in this study. Map from Fig. 1 of (Keyel et al., 2022) (public domain). Map base layers were derived from a combination of 2 public domain layers (US Census data, https://www.census.gov/geo/maps-data/data/tiger-line.html) and Natural Earth Administrative boundaries (https://www.naturalearthdata.com/downloads/50m-cultural-vectors/50m-admin-1-states-provinces).

**Fig. S3.**
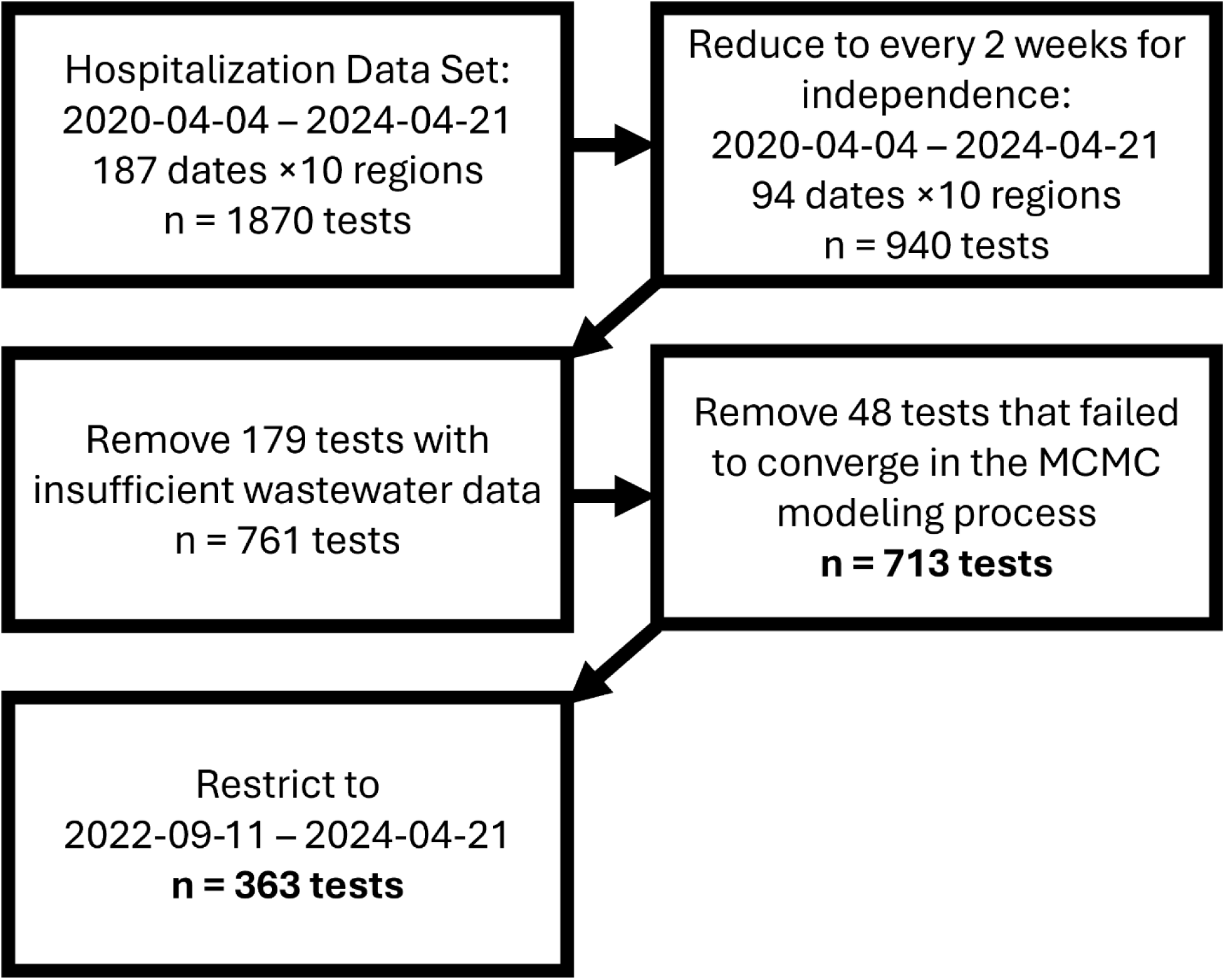
Flow chart describing the change in sample sizes from the initial data sets through the final analyses. Sample sizes decreased first as evaluations were conducted every 2 weeks to allow some measure of independence among observations. Observations were then screened for completeness of wastewater data. A subset of wwinference models failed to converge and those forecasts were excluded from the comparison. Finally, an additional date restriction was added to ensure the data set only included high statewide coverage.

**Fig. S4.**
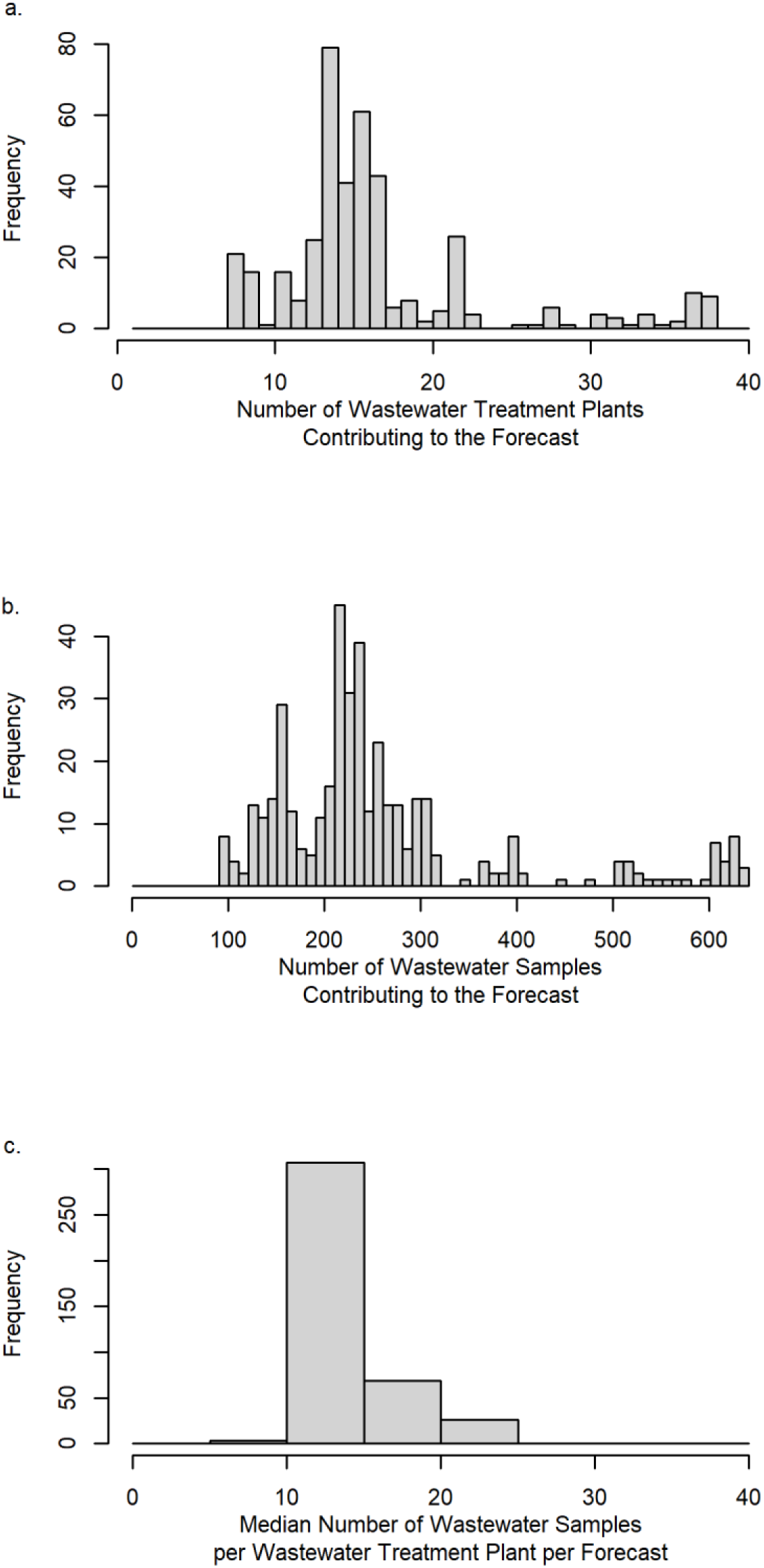
Summary information about a) the number of wastewater treatment plants, b) number of samples, and c) median number of samples per wastewater treatment plant that contributed to the forecasts.

**Table S1.**
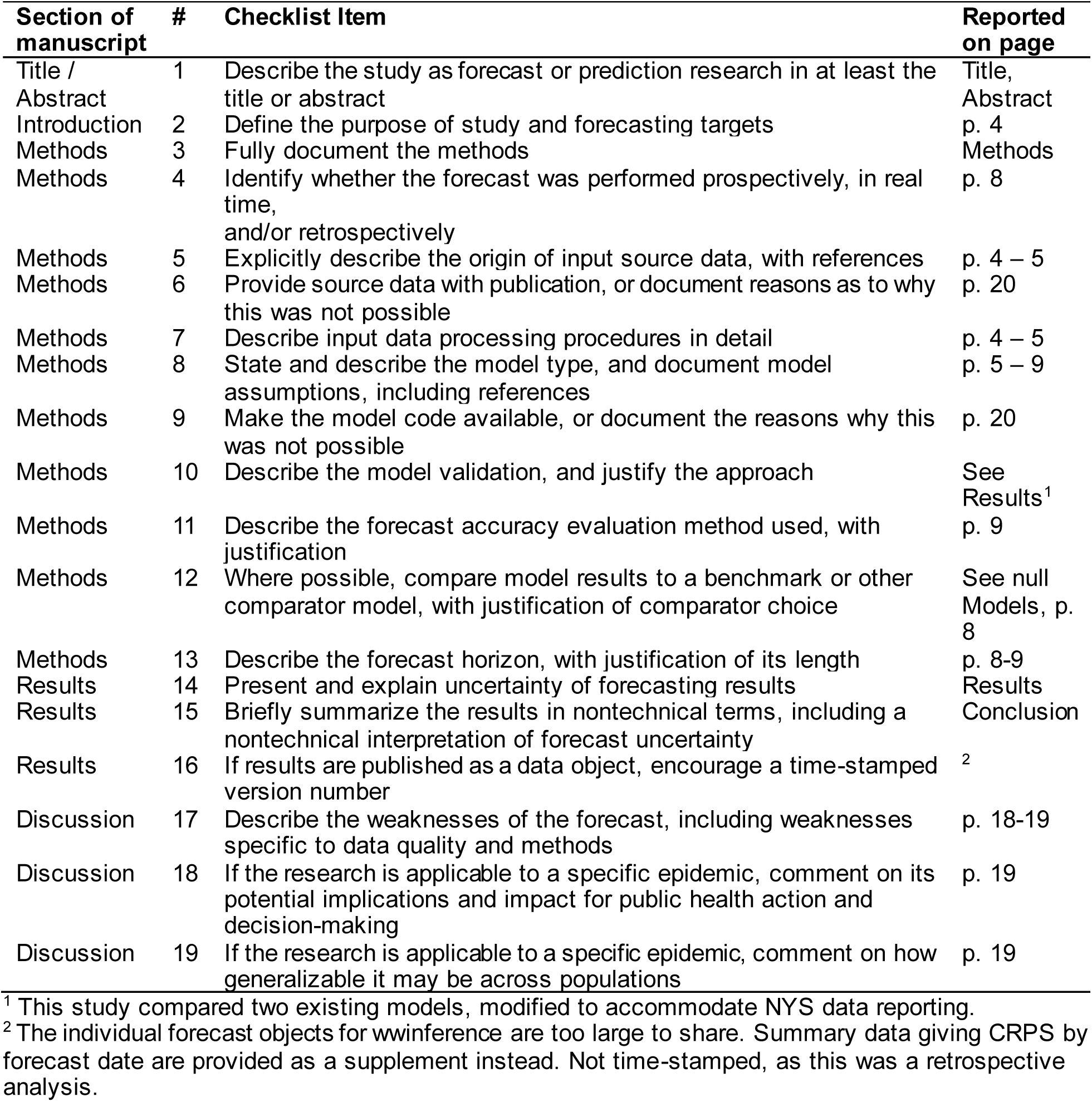
EPIFORGE 2020 checklist (Pollett et al., 2021)

**Table S2.**
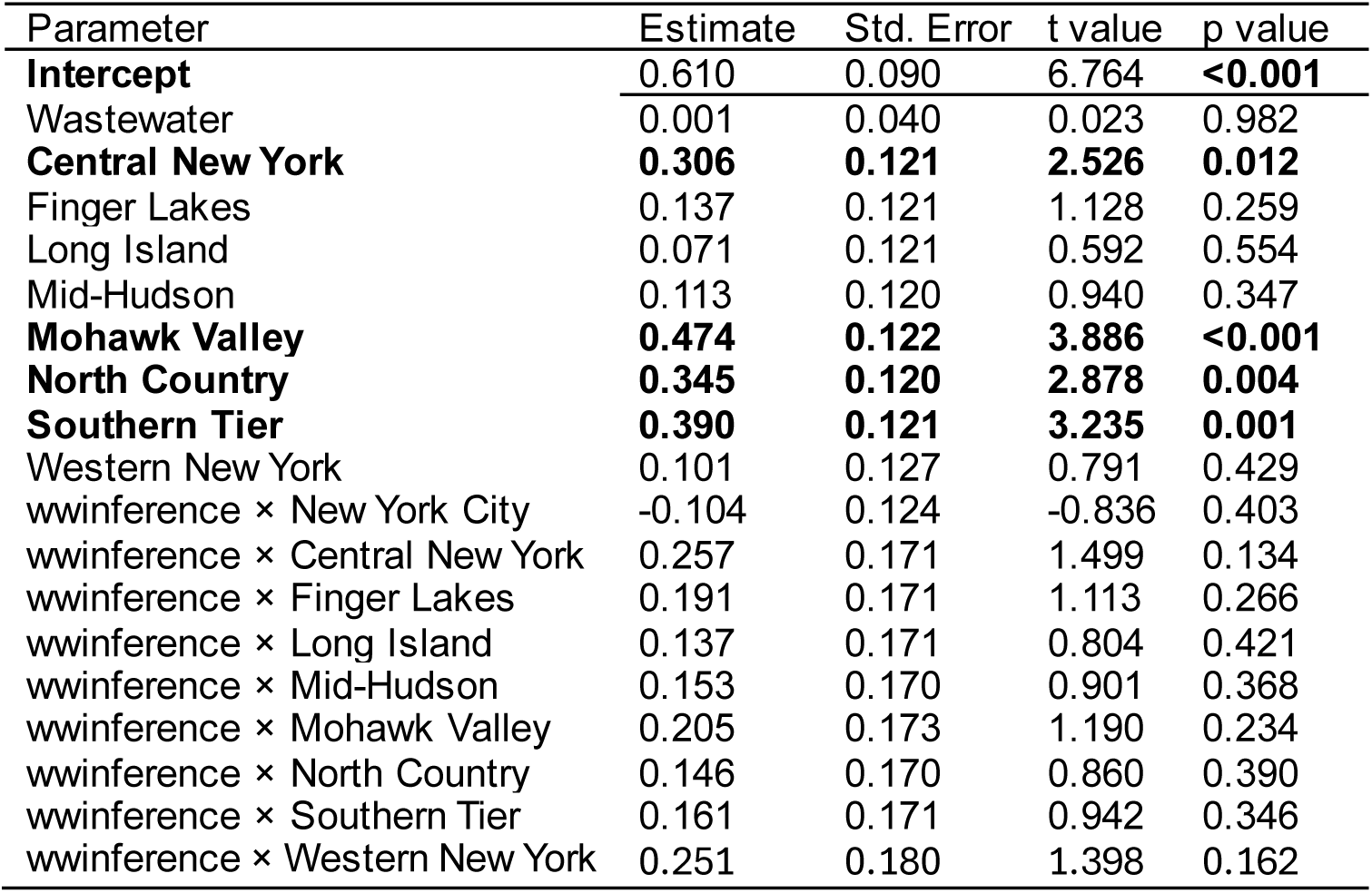
Natural scale regression results exploring differences in model performance by region. Reference levels were: GLMM (model), no wastewater, and New York City, the region with the lowest CRPS score on a natural scale. Bold indicates p < 0.05. N = 363 forecasts, null df = 1451, residual df = 1433.

| Parameter | Estimate | Std. Error | t value | p value |
| --- | --- | --- | --- | --- |
| **Intercept** | 0.610 | 0.090 | 6.764 | **<0.001** |
| Wastewater | 0.001 | 0.040 | 0.023 | 0.982 |
| **Central New York** | **0.306** | **0.121** | **2.526** | **0.012** |
| Finger Lakes | 0.137 | 0.121 | 1.128 | 0.259 |
| Long Island | 0.071 | 0.121 | 0.592 | 0.554 |
| Mid-Hudson | 0.113 | 0.120 | 0.940 | 0.347 |
| **Mohawk Valley** | **0.474** | **0.122** | **3.886** | **<0.001** |
| **North Country** | **0.345** | **0.120** | **2.878** | **0.004** |
| **Southern Tier** | **0.390** | **0.121** | **3.235** | **0.001** |
| Western New York | 0.101 | 0.127 | 0.791 | 0.429 |
| wwinference × New York City | -0.104 | 0.124 | -0.836 | 0.403 |
| wwinference × Central New York | 0.257 | 0.171 | 1.499 | 0.134 |
| wwinference × Finger Lakes | 0.191 | 0.171 | 1.113 | 0.266 |
| wwinference × Long Island | 0.137 | 0.171 | 0.804 | 0.421 |
| wwinference × Mid-Hudson | 0.153 | 0.170 | 0.901 | 0.368 |
| wwinference × Mohawk Valley | 0.205 | 0.173 | 1.190 | 0.234 |
| wwinference × North Country | 0.146 | 0.170 | 0.860 | 0.390 |
| wwinference × Southern Tier | 0.161 | 0.171 | 0.942 | 0.346 |
| wwinference × Western New York | 0.251 | 0.180 | 1.398 | 0.162 |

**Table S3.** **Log Scale** regression results exploring difference in model performance by region. Reference levels were: GLMM (model), no wastewater, and Finger Lakes, the region with the lowest CRPS score on a log scale. Bold indicates p < 0.05, italics and “.” indicate p <0.1 but ≥ 0.05. N = 363 forecasts, null df = 1451, residual df = 1433.

| Parameter | Estimate | Std. Error | t value | p value |
| --- | --- | --- | --- | --- |
| **Intercept** | **0.072** | **0.015** | **4.936** | **<0.001** |
| Wastewater | 0.000 | 0.007 | 0.060 | 0.952 |
| **Central New York** | **0.061** | **0.020** | **3.048** | **0.002** |
| Long Island | 0.021 | 0.020 | 1.049 | 0.294 |
| Mid-Hudson | 0.020 | 0.020 | 1.019 | 0.308 |
| **Mohawk Valley** | **0.151** | **0.020** | **7.513** | **<0.001** |
| *New York City* | *0.038* | *0.021* | *1.855* | *0.064* . |
| **North Country** | **0.131** | **0.020** | **6.610** | **<0.001** |
| **Southern Tier** | **0.125** | **0.020** | **6.282** | **<0.001** |
| Western New York | 0.033 | 0.021 | 1.592 | 0.111 |
| wwinference × Central New York | -0.004 | 0.028 | -0.158 | 0.875 |
| wwinference × Finger Lakes | 0.006 | 0.020 | 0.290 | 0.772 |
| wwinference × Long Island | -0.017 | 0.028 | -0.619 | 0.536 |
| wwinference × Mid-Hudson | -0.007 | 0.028 | -0.263 | 0.793 |
| wwinference × Mohawk Valley | 0.005 | 0.028 | 0.179 | 0.858 |
| *wwinference × New York City* | *-0.049* | *0.029* | *-1.681* | *0.093* . |
| wwinference × North Country | -0.010 | 0.028 | -0.346 | 0.729 |
| wwinference × Southern Tier | -0.006 | 0.028 | -0.215 | 0.830 |
| wwinference × Western New York | 0.012 | 0.030 | 0.418 | 0.676 |

**Table S4.**
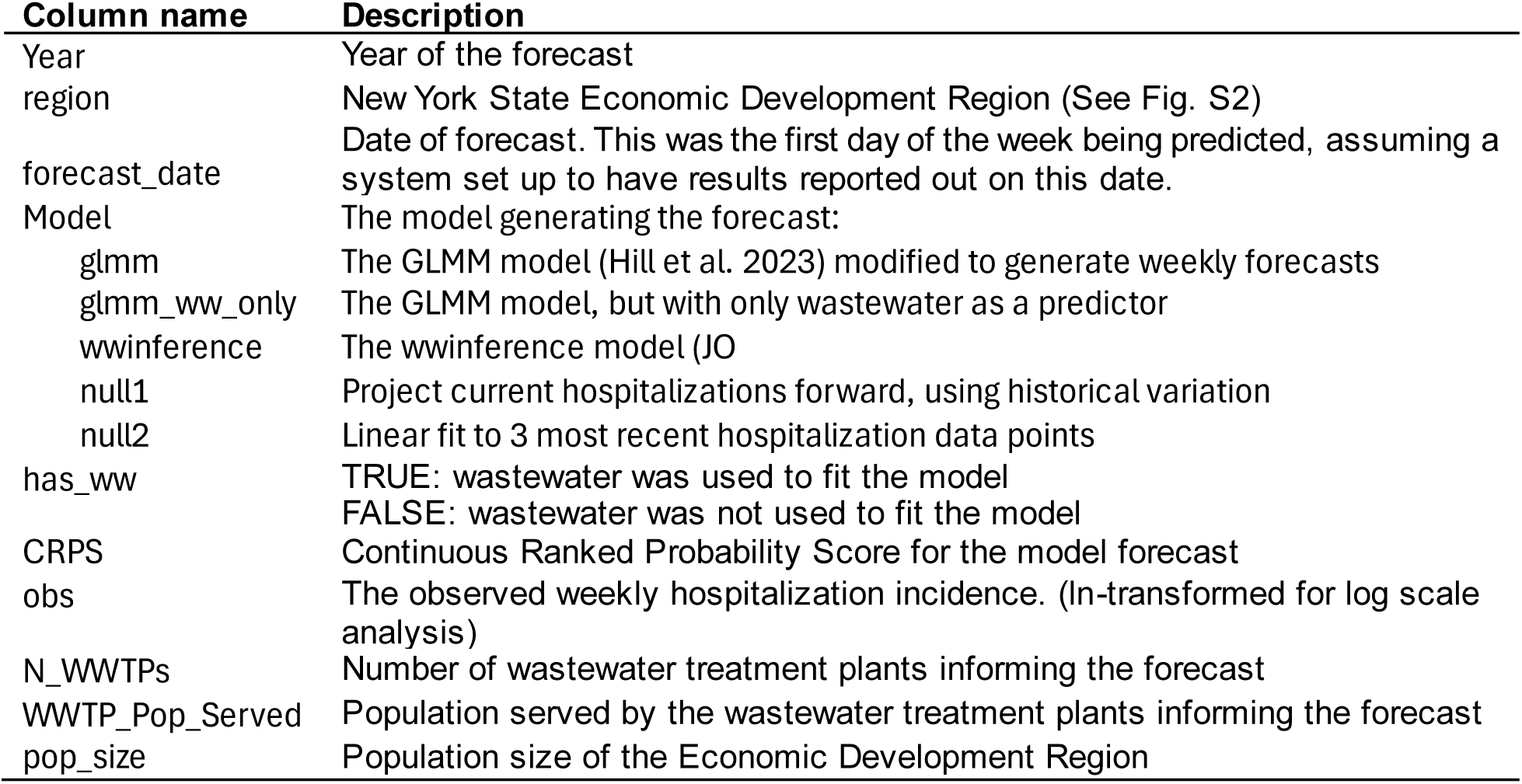
Data Dictionary for forecast evaluation objects (Supplement 2 and Supplement 3). The same column names were used for the natural scale and log scale results. Each file contains 2542 rows (including the header row) and 10 columns.

| Column name | Description |
| --- | --- |
| Year | Year of the forecast |
| region | New York State Economic Development Region (See Fig. S2) |
| forecast_date | Date of forecast. This was the first day of the week being predicted, assuming a system set up to have results reported out on this date. |
| Model | The model generating the forecast: |
| glmm | The GLMM model (Hill et al. 2023) modified to generate weekly forecasts |
| glmm_ww_only | The GLMM model, but with only wastewater as a predictor |
| wwinference | The wwinference model (JO) |
| null1 | Project current hospitalizations forward, using historical variation |
| null2 | Linear fit to 3 most recent hospitalization data points |
| has_ww | TRUE: wastewater was used to fit the model<br>FALSE: wastewater was not used to fit the model |
| CRPS | Continuous Ranked Probability Score for the model forecast |
| obs | The observed weekly hospitalization incidence. (ln-transformed for log scale analysis) |
| N_WWTPs | Number of wastewater treatment plants informing the forecast |
| WWTP_Pop_Served | Population served by the wastewater treatment plants informing the forecast |
| pop_size | Population size of the Economic Development Region |

**SUPPLEMENTS 2 and 3 ARE NOT INCLUDED IN THE PRE-PRINT**

